# An amplicon-based Illumina and nanopore sequencing workflow for Chikungunya virus West Africa genotype

**DOI:** 10.1101/2023.12.07.23299611

**Authors:** Idrissa Dieng, Mignane Ndiaye, Mouhamed Kane, Diamilatou Balde, Maimouna Mbanne, Seynabou Mbaye Ba Souna Diop, Safietou Sankhe, Moussa Dia, Madeleine Dieng, Serge Freddy Moukaha Doukanda, Ousmane Faye, Amadou Alpha Sall, Ndongo Dia, Gamou Fall, Oumar Faye, Moussa Moise Diagne

**Author notes:** These authors contributed equally to this article (Oumar Faye, Moussa Moise Diagne).

## Abstract

The Chikungunya virus, a global arbovirus, is currently causing a major outbreak in the Western African region, with the highest cases reported in Senegal and Burkina Faso. Recent molecular evolution analyses reveal that the strain responsible for the epidemic belongs to the West African genotype, with new mutations potentially impacting viral replication, antigenicity, and host adaptation. Real-time genomic monitoring is needed to track the virus’s spread in new regions. A scalable West African genotype amplicon-based Whole Genome Sequencing for multiple Next Generation Sequencing platforms has been developed to support genomic investigations and identify epidemiological links during the virus’s ongoing spread. This technology will help identify potential threats and support real-time genomic investigations in the ongoing spread of the virus.

## 1. Background

Chikungunya virus (CHIKV) is one of the most impactful arboviruses worldwide over the past twenty years. The spread of the virus in different continents has favored the emergence of different genotypes classified into West African (WA), East-Central-South-African (ECSA), Asian and Indian Ocean lineages (1).

CHIKV is identified in an increasing number of African countries (2). Given that most reported outbreaks in the continent are due to the ECSA group (3, 4, 5, 6), existing molecular characterization tools are mainly based on this genotype (7). However, most of the outbreaks in West Africa were due to the WA genotype (8). More recently, evidence of distribution area of this genotype has been reported with the virus detection in mosquitoes in China (9). At the time of writing, the Western African region is facing a great CHIKV outbreak with the highest numbers of cases ever reported in Senegal (10, 11) and Burkina Faso (12). Recent molecular evolution analyzes have shown that the strain responsible for the epidemic in Senegal belongs to the WA genotype and highlighted the emergence of new mutations with potential impact on viral replication, antigenicity and host adaptation (11). Moreover, the same study described transient amino acids substitutions. Thus, a real-time genomic monitoring is required following the progressive viral spread into new regions of Senegal (Public Health Direction in Institut Pasteur de Dakar) and potentially beyond through a reliable and accessible genomic characterization tool.

During the COVID-19 pandemic, the development of the amplicon-based Whole Genome Sequencing (WGS) allowed the acceleration of SARS-CoV-2 genomic surveillance in the world, even in the low-and-middle-income countries in Africa (13). In this work, we developed a scalable WA genotype CHIKV amplicon-based WGS for multiple NGS platforms in order to support the genomic investigations and identify epidemiological links during the ongoing spreading process of the virus.

## 2. Methods

### West African genotype primers design for Tiled Amplicon-Based Sequencing Systems

Based on all the available CHIKV sequences within the WA genotype and those from the ongoing Senegalese epidemic obtained using hybrid capture sequencing method (supplementary table S1), 400 bp primer pools (Supplementary table S2 tab 1) were designed using the ‘Primal Scheme’ tool (https://primalscheme.com/). To perform long sequencing reads, primer readjustments were carried out to constitute new pools capable of generating 1.2 kp amplicons with 400 bp of overlap between contiguous amplicons (Supplementary table S2 tab 2).

### Samples selection, nucleic acids extraction and RT-qPCR

Human sera from acute CHIKV positive cases collected in 2023 through both a CHIKV outbreak investigation in Kedougou (11) and the nationwide Syndromic Surveillance System in Senegal (14) were included. Sample selection was primarily based on viral load at diagnosis, with low to high CT values as the pattern.

A new extraction was done by the KingFisher Flex purification system (ThermoFisher, Darmstadt, Germany) using the MagMAX Viral/Pathogen Nucleic Acid Isolation Kit (ThermoFisher) following the manufacturer’s protocol. A CHIKV specific RT-qPCR (15) was performed on fresh RNA extracts before the sequencing experiments.

### Muliplex-PCR amplifications

Viral RNAs were reverse transcribed into cDNAs using LunaScript RT SuperMix (NEB #E3010) as previously described (16). Briefly, 8 μL template RNA and 2 μL LunaScript RT SuperMix (New England Biolabs #E3010) were mixed and put in the following thermal conditions: 25 °C for 10 min, followed by 50 °C for 10 min and 85 °C for 5 min. The CHIKV viral genome was amplified with either short (400 bp) and long-range (1.2 kp) PCR primers in two reaction pools using viral cDNA in the tiling PCR method. The synthesized cDNAs were template material for direct amplification to generate approximately 400 bp or 1.2 kp amplicons tiled along the genome using two systems of non-overlapping pools of WA-CHIKV targeting primers at 10 μM and Q5® High-Fidelity 2X Master Mix (New England Biolabs#M0494X) with the following thermal cycling protocol: 98 °C for 30 seconds, 35 cycles of: 95°C for 30 seconds, 65°C for 5 minutes, and a cooling step at 4°C.

### Illumina Sequencing

Sequencing libraries were synthesized by tagmentation of the 400 bp amplicons using the Illumina DNA Prep kit and the IDT® for Illumina PCR Unique Dual Indexes following manufacturer’s recommendations and as previously described (17). After a cleaning step with the Agencourt AMPure XP beads (Beckman Coulter, Indianapolis, IN), libraries were quantified using a Qubit 3.0 fluorometer (Invitrogen Inc., Waltham, MA, USA) for manual normalization before pooling in the sequencer. Cluster generation and sequencing were conducted in a Illumina iSeq100 instrument with iSeq 100 i1 Reagent v2 (300-cycle).

### Oxford Nanopore sequencing

Sequencing libraries preparation was carried out based on both 400 bp and 1.2 kp amplicons using the Oxford Nanopore Rapid Barcoding kit (SQK-RBK110.96) by attaching native barcodes (EXP-NBD114) and motor proteins to DNA molecules using a transposase as previously described (16). 16-plex library pools (for both 400 bp and 1.2 kp sequencing reads) were individually loaded into two different R9.4 flow cells (FLO-MIN106D) on a nanopore GridION sequencer with other parameters kept at their defaults.

### Sequencing data analysis

Raw data were collected and analyzed using guppy, raw reads in fastq format for both Illumina and nanopore approach were analyzed using Genome Detective tool (18) accessed on 24 November 2023. Sequences were aligned using MAFFT (19) and the alignment was run under the best model in IQ-TREE (20). The maximum-likelihood (ML) phylogenetic tree was visualized with metadata were performed using ggtree package implemented in R studio.

## 3. Results

### Chikungunya virus-West African genotype primers design for Amplicon-Based Sequencing

A multiplex primer system was designed using a WA-CHIKV reference genome, generating 36 oligonucleotide primer pairs for 400 bp tiled amplicons. The list of primers can be found in supplementary table S2. A long amplicon scheme was obtained by readjusting the first WA- CHIKV for a 1.2 kp long reads sequencing as shown by the supplementary table S2 (tab 1 and tab 2).

### Samples panel

Sixteen human sera from CHIKV positive cases in Senegal were included in the study based on viral load at diagnosis, with low to high CT values. RT-qPCR performed on the fresh RNA extracts showed consistent results between the ct values obtained during the first diagnostic and the ones from the new extraction except a sample previously tested positive on the field during an investigation in eastern Senegal that finally came negative in our study. These results are summarized in the table 2.

**Table 2.** Clinical samples panel for Chikungunya virus West Africa genotype amplicon-based sequencing.

| **Sample** | **Ct value upon**<br>**diagnostic** | **RNA ct value used during**<br>**the study** | **Area** |
| --- | --- | --- | --- |
| 417986 | 22.41 | 21.50 | Kedougou |
| 417905 | 23.03 | 27.04 | Kedougou |
| 418042 | 23.09 | 22.15 | Kedougou |
| 417854 | 23.87 | 23.60 | Kedougou |
| 418135 | 23.99 | 22.88 | Kedougou |
| 418017 | 24.24 | 24.26 | Kedougou |
| 417864 | 24.32 | 23.44 | Kedougou |
| 417861 | 24.36 | 24.16 | Kedougou |
| 417995 | 25.17 | 25.01 | Kedougou |
| 417824 | 25.29 | 27.53 | Kedougou |
| 418118 | 26.6 | 27.01 | Kedougou |
| 427924 | 27.17 | 21.04 | Agnam Civol |
| 417878 | 29.47 | 28.40 | Kedougou |
| 427972 | 30.03 | 29.01 | Agnam Civol |
| **427799** | 30.89 | NA | Tambacounda |
| **418013** | 32.67 | 33.45 | Kédougou |

### Short and long amplicon schemes comparison

Library preparations of the sample panel were undertaken as described above and simultaneously for both short and long amplicon schemes. cDNAs obtained by reverse transcription of the RNA extracts were amplified to get 400 bp and 1,200 bp amplicons. DNA concentrations measured by fluorimetry (Qubit) showed similar results between the two schemes (supplementary table S3).

Illumina-based sequencing was performed with the short (400 bp) amplicons while ONT approach was done using both 400 bp and 1.2 kp amplicons, as described above. Regardless of the sequencing technology, all schemes allowed to obtain complete or nearly complete genomes for all CHIKV positive RNAs (figure 1). Indeed, +80% horizontal coverage was systematically observed for all samples with ct value less than 30, and +90% for samples with the highest viral load (ct value ≤ 25). Overall, the ONT approach was quite similar in term of genome coverage than Illumina.

**Figure 1:**
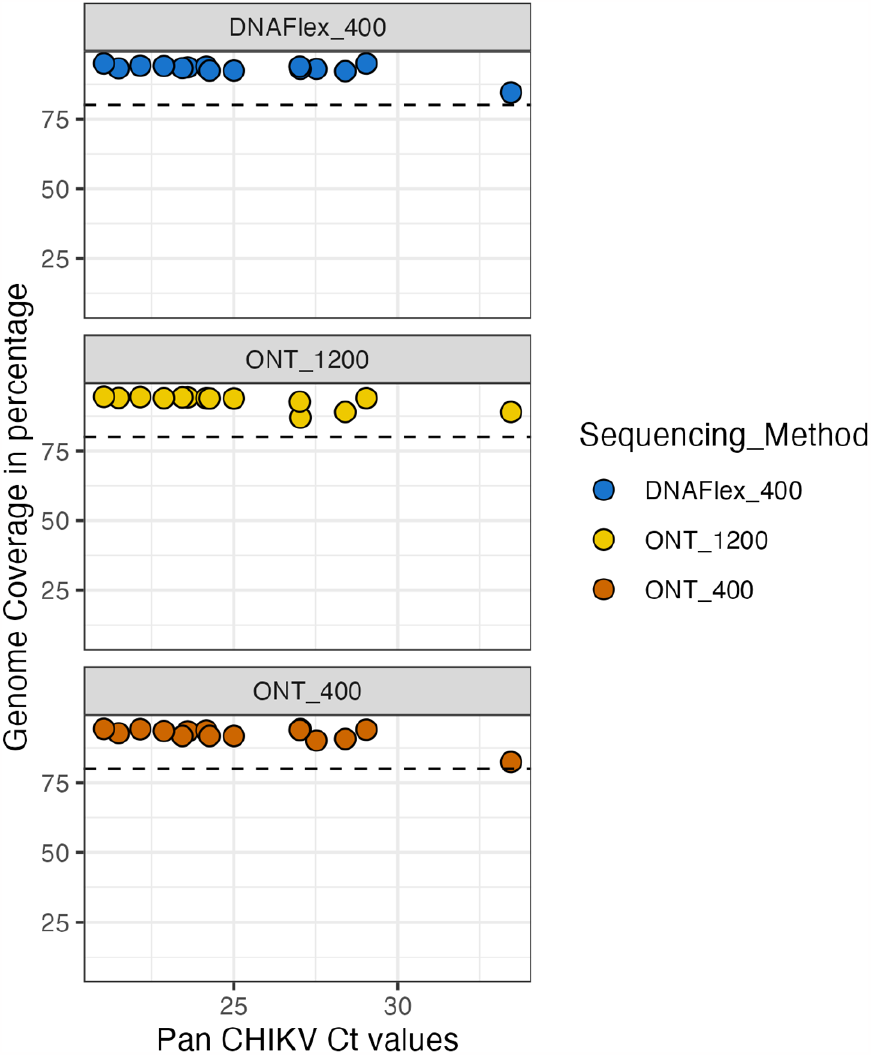
Relationship between the coverage of obtained CHIKV genomes and the pan CHIKV RT-qPCR ct values

Only samples 417824 was not properly sequenced by ONT using the long tiled amplicons while the complete cds was obtained using the short scheme either by Illumina and ONT. The sample 418013 with the lowest viral load (table 1) was poorly covered with a maximum of half the genome obtained by the long amplicon scheme approach on ONT (supplementary figure S1). In general, the number of reads generated by Illumina sequencing was greater than ONT. Among the ONT runs, the long amplicon scheme most often had a higher overall number of reads than the short-PCR t (figure 2).

**Figure 2.**
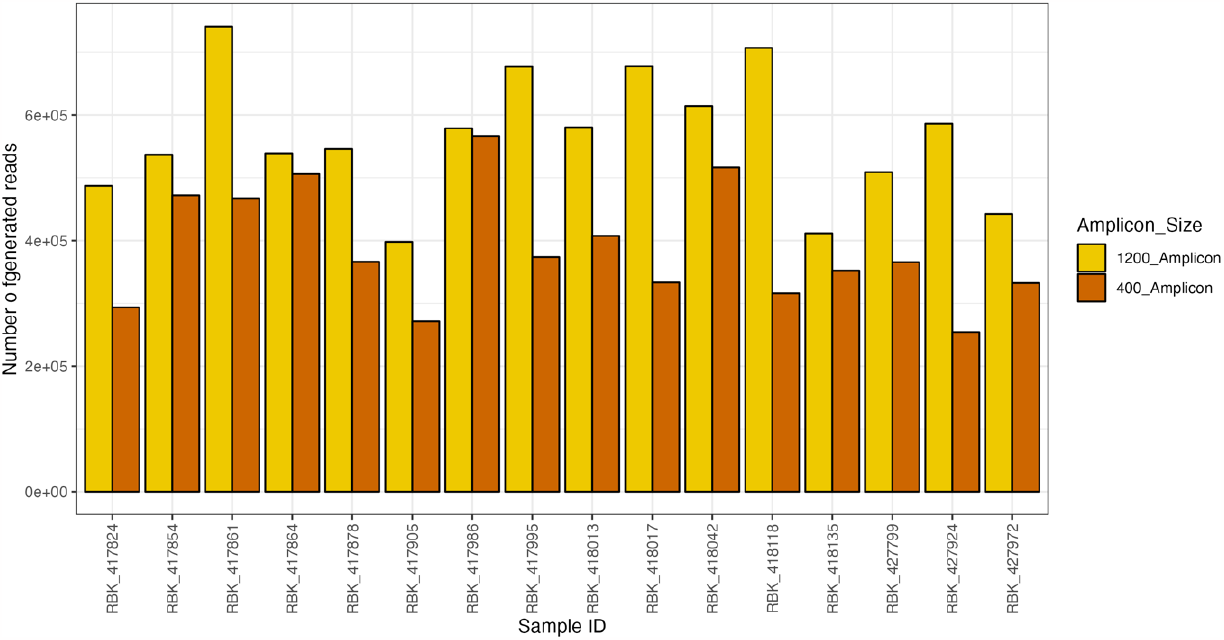
Number of generated reads per clinical sample for both short and long amplicon scheme in Oxford Nanopore Technology.

However, excepted two or three samples, the number of mapped reads to the CHIKV reference genome was quite similar between both schemes with ONT while the short tiled PCR method allowed to generate more specific data though Illumina. Only the sample that came back negative after re-extraction (sample 427799) did not have reads mapping to CHIKV whatever the sequencing method or the scheme (figure 3). Moreover, sample 427972 gave much less reads from the long amplicon scheme in ONT compared to the two other methods.

**Figure 3:**
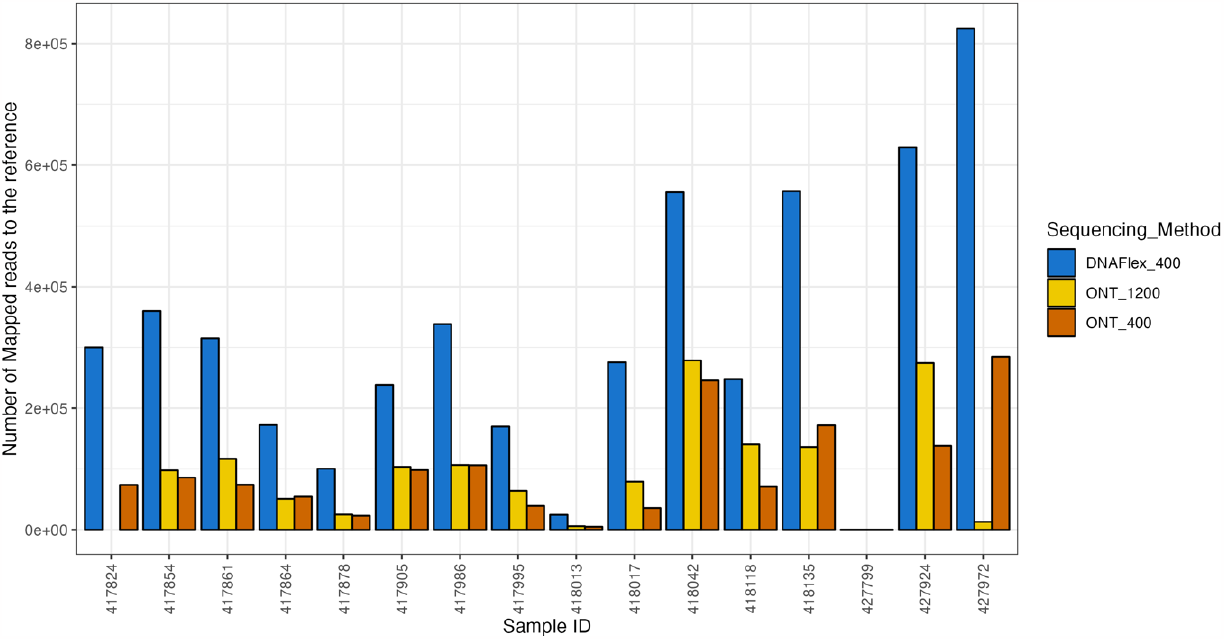
Number of mapped reads against used CHIKV reference for the different sequencing platforms and methods

A ML tree was built using sequence data obtained from fifteen clinical samples to evaluate the consistency of different schemes and approaches in terms of genomic information. The analysis showed that in most cases (+93%), sequences generated from the same sample group together regardless of the sequencing method used, the sample with the lowest viral load also being included (figure 4). Only one sample (417986) showed great divergence between the three sequences obtained with the different methods despite the generation of a valuable number of reads mapping with CHIKV for both short and long amplicon schemes in Illumina or ONT sequencers (figure 3).

**Figure 4.**
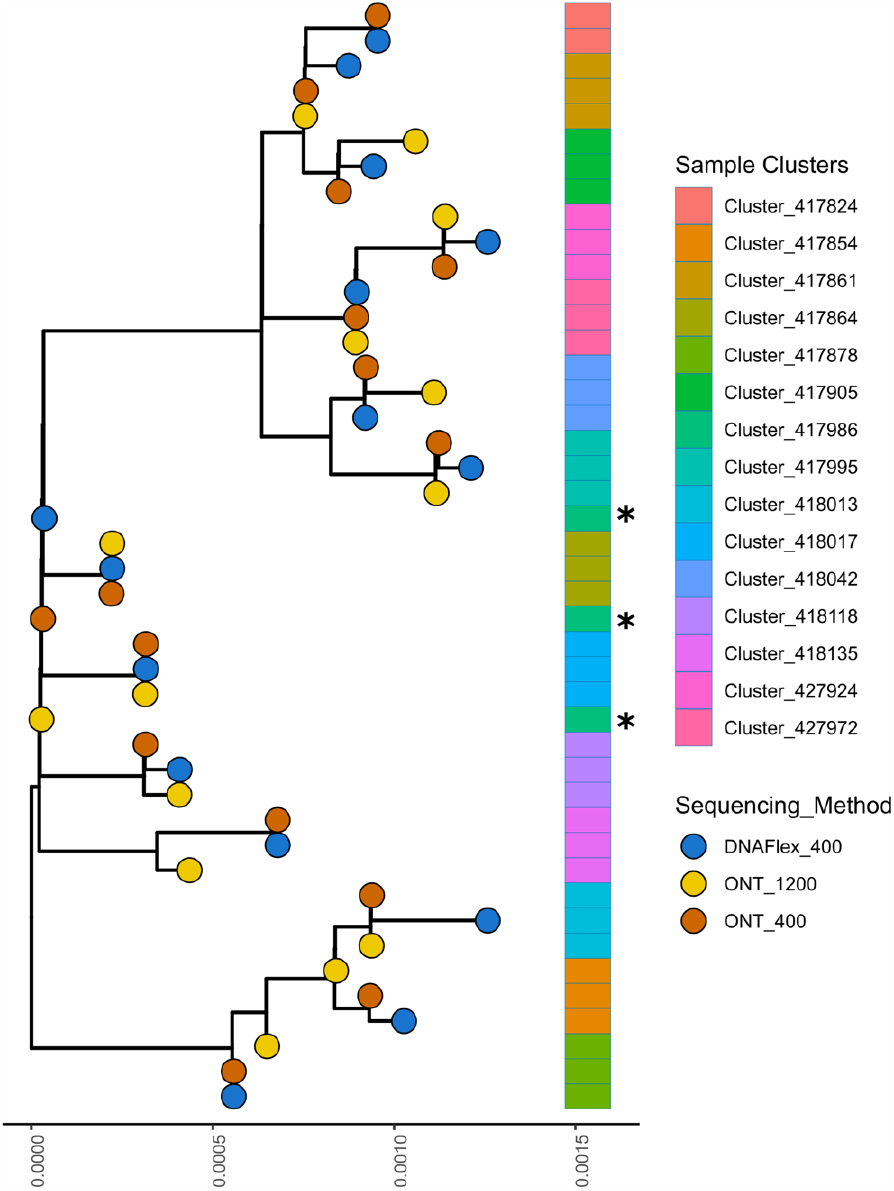
Phylogenic tree of the sequences obtained from the clinical samples sets by the different amplicon schemes and sequencing methods.

## Discussion

In this study, we aimed to develop a scalable CHIKV amplicon-based WGS to fill the gap in existing sequencing tools for monitoring the epidemiological and molecular evolution of CHIKV. Indeed, to our knowledge, most of the available methods are based on the most widespread ECSA genotype.

Although emergence caused by the WA genotype are still localized, we are now seeing increasingly explosive outbreaks such as the one underway in Senegal (11). We designed a short amplicon scheme for WA-CHIKV WGS. The same primers were also used to develop a long amplicon scheme. The validation of the two schemes were done on a panel of clinical samples diagnosed positive to acute CHIKV infection. Both schemes were used for ONT sequencing while Illumina sequencing was performed only with the short tiled PCR method. The validation process’s data demonstrated consistency between short and long amplicon schemes regardless of the sequencing technologies. Near-complete or complete genome sequences were obtained from almost the entire panel of CHIKV-positive clinical samples.

If Illumina approach offered the greatest amount of data, sequencing via the long amplicon scheme has shown slightly better results when we talk about ONT sequencing. Moreover, a good consistency in term of CHIKV sequences was globally observed from one sequencing method to another one, highlighting that sequencing by ONT with either the short or long WA-CHIKV amplicon schemes gave results similar to the gold standard Illumina sequencing by synthesis on short amplicons. Nanopore technology’s sequencing accuracy is often considered a weakness, but it was shown that a 10x sequencing depth cutoff yielded a consensus sequence accuracy of about 99%, similar to Illumina sequences (21). The solutions proposed in this work greatly exceed this depth cutoff.

Thus, in addition to Illumina-based sequencing, we describe systems adapted to ONT for equally reliable results. The ONT sequencers are portable, cost-effective and highly scalable NGS platforms that streamline sequencing workflow and reduces turnaround times. This work provides versatile WA-CHIKV WGS schemes of interest for real-time molecular epidemiology and evolutionary studies.

## Data Availability

All data produced in the present study are available upon reasonable request to the authors

## Acknowledgments

We thank the Collaborating Center for Arboviruses and Hemorrhagic Fever viruses in the Virology Department of Institut Pasteur de Dakar for their critical role in the diagnostic process.

## Informed consent

Clinical samples in this study were collected on CHIKV positive human samples according to the guidelines of the Declaration of Helsinki, and approved by the Senegalese national ethics committee for health research (protocol code SEN20/08 approved on 6 April 2020).

## References

1. de Souza WM, de Lima STS, Simões Mello LM, Candido DS, Buss L, Whittaker C, Claro IM, Chandradeva N, Granja F, de Jesus R, Lemos PS, Toledo-Teixeira DA, Barbosa PP, Firmino ACL, Amorim MR, Duarte LMF, Pessoa IB Jr, Forato J, Vasconcelos IL, Maximo ACBM, Araújo ELL, Perdigão Mello L, Sabino EC, Proença-Módena JL, Faria NR, Weaver SC. 2023. Spatiotemporal dynamics and recurrence of chikungunya virus in Brazil: an epidemiological study. Lancet Micr 4(5):e319–e329.

2. World Health Organization. Chikungunya. Available at: https://www.afro.who.int/health-topics/chikungunya

3. Yonga MG, Yandai FH, Sadeuh-Mba S, Abdallah AH, Ouapi D, Gamougam K, Abanda NN, Endengue-Zanga MC, Demanou M, Njouom R. 2022. Molecular characterization of chikungunya virus from the first cluster of patients during the 2020 outbreak in Chad. Arch Virol 167(5):1301–1305.

4. Agbodzi B, Yousseu FBS, Simo FBN, Kumordjie S, Yeboah C, Mosore MT, Bentil RE, Prieto K, Colston SM, Attram N, Nimo-Paintsil S, Fox AT, Bonney JHK, Ampofo W, Coatsworth HG, Dinglasan RR, Wolfe DM, Wiley MR, Demanou M, Letizia AG. 2021. Chikungunya viruses containing the A226V mutation detected retrospectively in Cameroon form a new geographical subclade. Int J Infect Dis 113:65–73.

5. Chinedu Eneh S, Uwishema O, Nazir A, El Jurdi E, Faith Olanrewaju O, Abbass Z, Mustapha Jolayemi M, Mina N, Kseiry L, Onyeaka H. Chikungunya outbreak in Africa: a review of the literature. 2023. Ann Med Surg (Lond) 19;85(7):3545–3552.

6. Russo G, Subissi L, Rezza G. Chikungunya fever in Africa: a systematic review. 2020. Pathog Glob Health 114(3):136–144.

7. de Souza LM, de Oliveira ID, Sales FCS, da Costa AC, Campos KR, Abbud A, Guerra JM, Dos Santos Cirqueira Borges C, Takahashi Cpfj, de Araújo LJT. Technical comparison of MinIon and Illumina technologies for genotyping Chikungunya virus in clinical samples. 2023. J Genet Eng Biotechnol 29;21(1):88.

8. Sow A, Faye O, Diallo M, Diallo D, Chen R, Faye O, Diagne CT, Guerbois M, Weidmann M, Ndiaye Y, Senghor CS, Faye A, Diop OM, Sadio B, Ndiaye O, Watts D, Hanley KA, Dia AT, Malvy D, Weaver SC, Sall AA. 2017. Chikungunya Outbreak in Kedougou, Southeastern Senegal in 2009-2010. Open Forum Infect Dis 2;5(1):ofx259.

9. Li N, Peng C, Yuan Y, Hao Y, Ma W, Xiao P. 2023. A new cluster of chikungunya virus West Africa genotype isolated from Aedes albopictus in China. J Infect. 87(3):e48–e50.

10. Ministry of health and social action in Senegal website. Available at: https://www.sante.gouv.sn

11. Dieng I, Sadio BD, Gaye A, Sagne SN, Ndione MHD, Kane M, Diallo MK, Sow B, Sankhe S, Diallo A, Dieng M, Doukanda SFM, Mbanne M, Diop SMBS, Balde D, Ndiaye M, Sow KD, Diarra M, Sam A, Mbaye A, Diallo B, Sall Y, Faye O, Diop B, So A, Sall AA, Loucoubar C, Dia N, Faye Oum, Diallo D, Fall G, Weaver SC, Barry MA, Diallo M, Diagne MM. 2023. Genomic characterization of a reemerging Chikungunya outbreak in Kedougou, Southeastern Senegal, 2023. [preprint]. Available at: 10.13140/RG.2.2.32038.50249

12. Center for Disease control and Prevention. Chikungunya in Burkina Faso. Available at: https://www.nc.cdc.gov/travel/notices/level2/chikungunya-burkina-faso

13. Tegally H, San JE, Cotten M, Moir M, Tegomoh B, Mboowa G, Martin DP, Baxter C, Lambisia AW, Diallo A, Amoako DG, Diagne MM, Sisay A, Zekri AN, Gueye AS, Sangare AK, Ouedraogo AS, Sow A, Musa AO, Sesay AK, …Wilkinson E. 2022. The evolving SARS-CoV-2 epidemic in Africa: Insights from rapidly expanding genomic surveillance. Science 7;378(6615):eabq5358.

14. Diagne MM, Ndione MHD, Gaye A, Barry MA, Diallo D, Diallo A, Mwakibete LL, Diop M, Ndiaye EH, Ahyong V, Diouf B, Mhamadi M, Diagne CT, Danfakha F, Diop B, Faye O, Loucoubar C, Fall G, Tato CM, Sall AA, Weaver SC, Diallo M, Faye O. 2021. Yellow Fever Outbreak in Eastern Senegal, 2020-2021. Viruses. 28;13(8):1475.

15. Diallo D, Sall AA, Buenemann M, Chen R, Faye O, Diagne CT, Faye O, Ba Y, Dia I, Watts D, Weaver SC, Hanley KA, Diallo M. 2012. Landscape ecology of sylvatic chikungunya virus and mosquito vectors in southeastern Senegal. PLoS Negl Trop Dis 6(6):e1649.

16. Freed NE, Vlková M, Faisal MB, Silander OK. 2020. Rapid and inexpensive whole-genome sequencing of SARS-CoV-2 using 1200 bp tiled amplicons and Oxford Nanopore Rapid Barcoding. Biol Methods Protoc 18;5(1):bpaa014.

17. Diagne MM, Ndione MHD, Mencattelli G, Diallo A, Ndiaye EH, Di Domenico M, Diallo D, Kane M, Curini V, Top NM, Marcacci M, Mbanne M, Ancora M, Secondini B, Di Lollo V, Teodori L, Leone A, Puglia I, Gaye A, Sall AA, Loucoubar C, Rosà R, Diallo M, Monaco F, Faye O, Cammà C, Rizzoli A, Savini G, Faye O. 2023. Novel Amplicon-Based Sequencing Approach to West Nile Virus. Viruses. 27;15(6):1261. doi: 10.3390/v15061261.

18. Vilsker M, Moosa Y, Nooij S, Fonseca V, Ghysens Y, Dumon K, Pauwels R, Alcantara LC, Vanden Eynden E, Vandamme AM, Deforche K, de Oliveira T. 2019. Genome Detective: an automated system for virus identification from high-throughput sequencing data. Bioinformatics. 1;35(5):871–873.

19. Katoh K, Misawa K, Kuma K, Miyata T. MAFFT: a novel method for rapid multiple sequence alignment based on fast Fourier transform. 2002. Nucleic Acids Res. 15;30(14):3059–66. doi: 10.1093/nar/gkf436.

20. Nguyen LT, Schmidt HA, von Haeseler A, Minh BQ. IQ-TREE: a fast and effective stochastic algorithm for estimating maximum-likelihood phylogenies. 2015. Mol Biol Evol. 32(1):268–74.

21. Charre C, Ginevra C, Sabatier M, Regue H, Destras G, Brun S, Burfin G, Scholtes C, Morfin F, Valette M, Lina B, Bal A, Josset L. 2020. Evaluation of NGS-based approaches for SARS-CoV-2 whole genome characterisation. Virus Evol. 5;6(2):veaa075.

