## Supplementary figures and images for "An amplicon-based Illumina and nanopore sequencing workflow for Chikungunya virus West Africa genotype"

### Supplementary figure S1.pdf

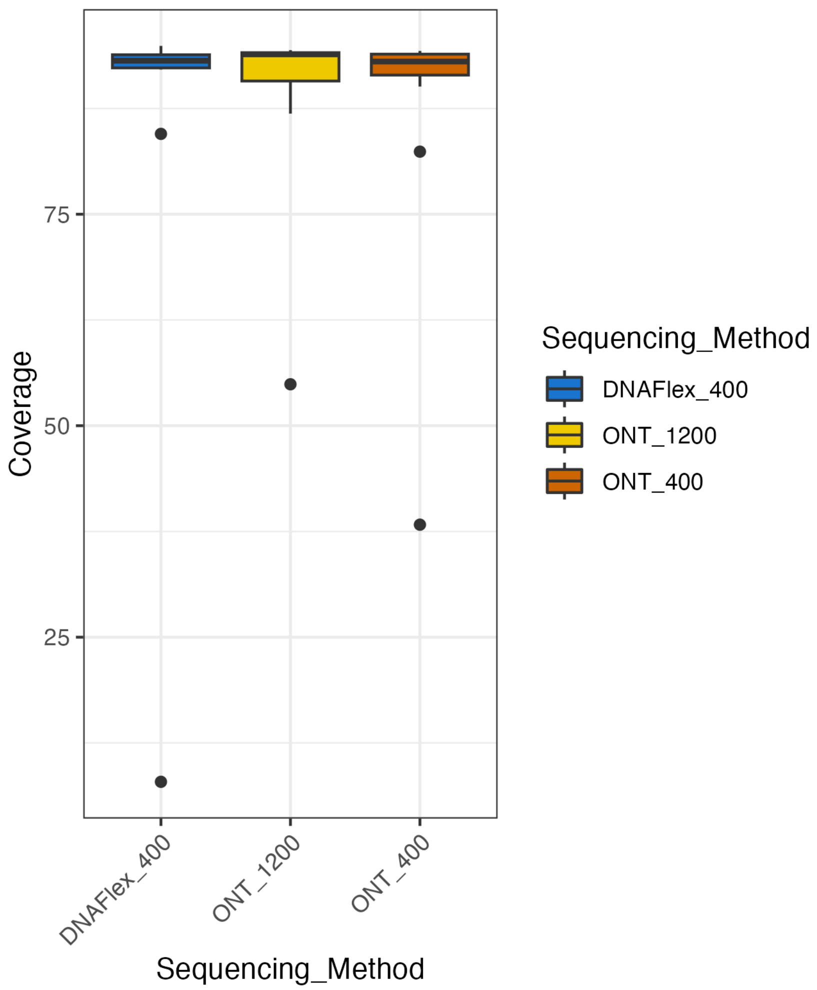

### Supplementary figure S1.png

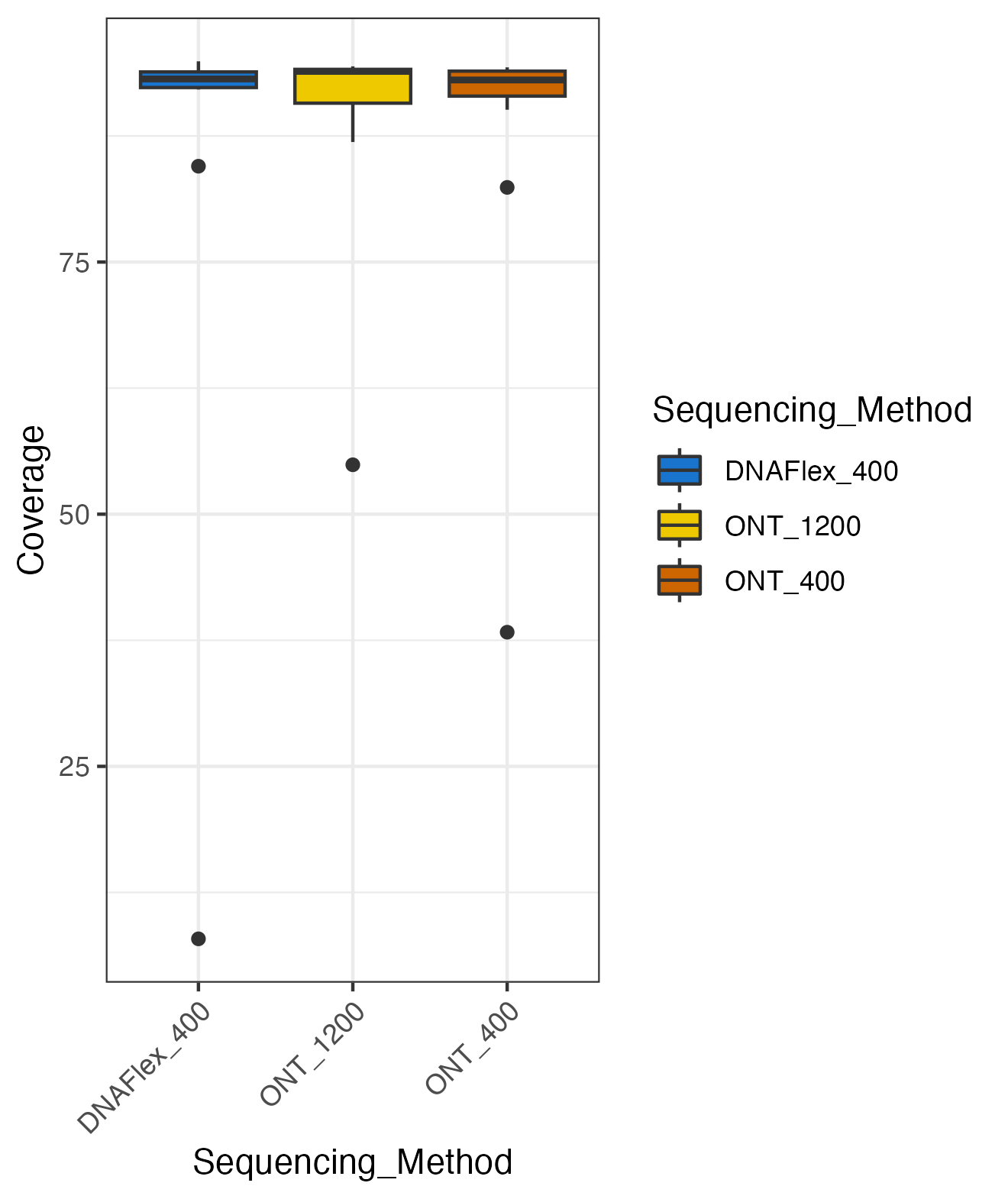
